# Impact of COVID-19 pandemic on Black, Asian and Minority Ethnic (BAME) communities: a qualitative study on the perspectives of BAME community leaders

**DOI:** 10.1101/2021.03.03.21252286

**Authors:** Fesani Mahmood, Dev Acharya, Kanta Kumar, Vibhu Paudyal

## Abstract

**Objectives:** The aim of this study was to explore the perspectives of BAME community leaders in relation to - the impact of the COVID-19 pandemic on their communities; and BAME community’s perception, understanding and adherence to Government guidelines on COVID-19 public health measures.

**Design:** A phenomenological approach was adopted using qualitative semi-structured interviews.

**Settings:** Community organisations and places of worships in the West Midlands region of England.

**Participants:** Community leaders were recruited through organisations representing BAME communities and religious places of worship.

**Results:** A total of 19 participants took part. Participants alluded to historical and structural differences for the observed disparities in COVID-19 morbidity and mortality. Many struggled with lockdown measures which impeded cultural and religious gatherings that were deemed to be integral to the community. Cultural and social practices led to many suffering on their own as discussion of mental health was still deemed a taboo within many communities. Many expressed their community’s reluctance to report symptoms for the fear of financial and physical health implications. They reported increase in hate crime which was deemed to be exacerbated due to perceived insensitive messaging from authority officials and historical structural biases. Access and adherence to government guidelines was an issue for many due to language and digital barriers. Reinforcement from trusted community and religious leaders encouraged adherence. Points of support such as food banks were vital in ensuring essential supplies during the pandemic. Many could not afford masks and sanitisers.

**Conclusion:** The study highlights the perceived impact of COVID-19 pandemic on BAME communities. Government agencies and public health agencies need to integrate with the community, and community leaders to penetrate the key messages and deliver targeted yet sensitive public health advice which incorporates cultural and religious practices. Addressing route cause of disparities is imperative to mitigate current and future pandemics.

**Strengths and limitations of this study:**

- To our knowledge, this is the first study in England to investigate the understanding of risk and impact of COVID-19 using the perspectives of BAME community leaders.
- Participants represented diverse BAME community organisations and places of worship.
- Participant recruitment was limited to one of the seven regions within England with the highest proportion of BAME populations.
- Results may not be generalizable to any BAME communities not represented in the data.

## INTRODUCTION

Coronavirus disease (COVID-19) was declared a global pandemic in March 2020, with over 120,000 deaths from the virus in the UK as of February 2021.^1^ There was early recognition that Black, Asian and Minority Ethnic (BAME) groups in the UK were disproportionately affected, ^2,3^ which came to public attention when the first ten doctors who had died from COVID-19 were of BAME origin.^4^ During the first wave in April 2020, approximately 35% of almost 2,000 intensive care patients for COVID-19 in England, Wales and Northern Ireland were non-White.^5^ However, BAME groups only constitute 13% of the UK’s population.^6^ Recent estimates suggest that Chinese, Indian, Pakistani, other Asian, Black Caribbean and other Black ethnicity had between 10-50% higher mortality risk compared to the White British population.^7^

Historically, health inequalities have been a concern for BAME groups; for example, disparity has been observed through higher prevalence of type 2 diabetes and cardiovascular disease in South Asian communities.^8^ The concept is defined as “avoidable and unfair differences” shaped by their surrounding circumstances, namely wider determinants of health.^9^ Factors such as socio-economic status, cultural practices and environmental conditions –which are largely influenced by structural biases based on ethnicity, collectively drive inequalities. Whilst literature reinforces the understanding that BAME groups are disproportionately affected by COVID-19, there is a lack of research that aims to understand the disparity from the perspectives of BAME communities.

Urgency of further study into the association between ethnicity and COVID-19 was highlighted in research and policy domains in early 2020. Data from the Office for National Statistics (ONS) proposed that existing co-morbidities in BAME COVID-19 patients could have contributed towards the disparity.^10^ However, the debate later incorporated wider social and structural disparities. Factors such as deprivation, living conditions, nature of employment were linked to higher morbidity and mortality in BAME populations.^11^

In addition to the disparities in morbidity and mortality directly as a result of COVID-19, anecdotal media reports have stated that BAME communities have been under protected, stigmatised during this pandemic.^12^ Currently there is a sparse literature exploring the understanding of BAME groups on how they, themselves, perceive their disparity in COVID-19. To mitigate the pandemic and its impact, diverse information were disseminated through social and broadcast media by the UK Government and public health organisations. Slogans such as ‘Stay Home, Protect the NHS, Save Lives’ endeavoured to support the first national lockdown in the UK. During this period, non-essential businesses, community organisations and places of worship, which are integral to many BAME communities, were inaccessible.^13^ Downloadable translations of key documents, including posters for COVID-19 symptoms were made available in England through Public Health England in eleven different languages.^14^ However, BAME communities’ understanding of pandemic-related communication from the Government and public health organisations has not yet been investigated.

The aim of this study was to explore the perspectives of BAME community leaders in relation to: the impact of the COVID-19 pandemic on their communities; and BAME community’s perception, understanding and adherence to Government guidelines on COVID-19 public health measures.

## METHODS

### Design

A phenomenological approach using qualitative study design was adopted.^15^

### Study population, sampling strategy and recruitment

Leader or representatives of the organisations, businesses and places of worship serving a predominantly BAME community in the West Midlands region of England, a region that suffered most BAME related hospital admissions and mortality, were searched online, then invited via email or telephone. Organisations were identified through internet, social media search and acquaintance of the research team (all representing BAME communities). Those expressing interest were emailed a Participant Information Sheet and Consent Form. Additional recruitment was made through snowball sampling. The ethnic groups of interest (Electronic supplementary material 1) were of those recommended by the UK Government.^16^

### Ethical approval and consent

Ethical approval was obtained from the University of Birmingham School of Pharmacy Ethics Committee (reference number UoB/SoP/2020-64). Informed consent was received from all participants.

### Data collection material and methods

An interview topic guide (Electronic supplementary material 2) was developed with 19 open-ended questions on three key areas regarding BAME community’s: (1) understanding of acquiring COVID-19 risk and disparity in health outcomes; (2) beliefs and perspectives relating to COVID-19; and (3) understanding and adherence to Government-issued guidance and public health measures on COVID-19. Probes elicited a comprehensive insight from participants. To test face and content validity, the research team developed questions based on existing literature and a pilot interview was conducted with a BAME community member which ensured clarity of the questions. No changes had to be made to the topic guide.

Participants were recruited and interviewed between October - November 2020 for approximately 45 minutes over Zoom or telephone. Relevant demographic information was collected prior to the interview. Interviews were audio-recorded using the recording function on Zoom or a digital voice recorder, respectively.

### Data processing and analysis

Recordings were transcribed verbatim into Microsoft Word. Data was anonymised to remove any identifiable information. Transcripts were exported onto Microsoft Excel and then thematically analysed by two researchers (FM and VP) using framework technique.^17–19^ The initial coding was reviewed between the research team through analysis of first two transcripts before an agreed version that could be applied to the rest of the transcripts. New themes were as they emerged during the subsequent analysis of other transcripts.

Reporting was conducted to comply with the COREQ guideline^20^ and checklist (electronic supplement material 3).

#### Patient and public involvement

Apart from members of public’s participation in the research as study subjects, no other PPI activities were conducted for this research due to time and resource constraints.

## RESULTS

A total of 19 participants from various community leadership roles and BAME representations took part (table 1). Four overarching themes and 11 subthemes were identified (tables 2–5). Themes related to a) perceived impact of COVID-19 and lockdown on wellbeing b) understanding of risk and disparity in health outcomes for COVID-19; c) perception, understanding and adherence to Government guidance in relation to COVID-19; d) accessibility and use of community services, and other points of support, during the pandemic. Narrative summaries of each theme are presented below and illustrative quotes are presented in tables 2, 3, 4 and 5 dedicated to each theme.

**Table 1:** Demographics of recruited study participants.

|  | **Participant community role** | **Broad ethnic group of community** |
| --- | --- | --- |
| P1 | Community service provider | Asian/British Asian |
| P2 | School teacher | Asian/British Asian |
| P3 | Pharmacist | Asian/British Asian |
| P4 | Church staff | Black/African/Caribbean/British Black |
| P5 | Mosque imam | Asian/British Asian |
| P6 | Radio presenter | Black African/British Black African |
| P7 | Community service provider | Asian/British Asian |
| P8 | Community service provider | Asian/British Asian |
| P9 | Supermarket manager | Asian/British Asian |
| P10 | Mosque imam | Various BAME groups |
| P11 | Church leader | Black/African/Caribbean/British Black |
| P12 | Community service provider | Yemeni/British Yemeni |
| P13 | Temple manager | Asian/British Asian |
| P14 | Community service provider | Various BAME groups |
| P15 | Church staff | Black/African/Caribbean/British Black |
| P16 | Community service provider staff | Black/African/Caribbean/British Black |
| P17 | School teacher | Asian/British Asian |
| P18 | Community service provider | Black/African/Caribbean/British Black |
| P19 | Community service provider | Black/African/Caribbean/British Black/White and Black Caribbean/White and Black African |

**Table 2.** Perceived impact of COVID-19 and lockdown on wellbeing.

| Thematic subtheme | Thematic code | Illustrative quotes |
| --- | --- | --- |
| Mental health impact and psychological wellbeing | Isolation | *"in the Asian community... from going to mixing with lots of family then all of a sudden you might not, that's very hard for people"*<br>- P17, Asian/British Asian, school teacher |
|  | Emotions and emotional fatigue | *"the younger ladies... struggled as well..., the kids were at home and they were saying they're driving them insane, having to do the home-schooling... all the cooking, cleaning... school gives that break... They found it initially... very enjoyable being with the family but with a longer period of time it does get a bit repetitive, kids become ratty, they become...very frustrated... I remember one parent saying to me... she found it really exhausting."*<br>- P7, Asian/British Asian, community service provider |
|  | Taboo | *"mental health has a very big stigma in my people..., they don't talk about mental health, it's a cultural thing. Mental health, they'll tell you, oh my God, audhu billahi minashaitan [I seek refuge in God from the outcast Satan], ... what you're talking about, you don't believe in Allah [God]? ... why are you saying you have a mental health? People will stamp on you... if you say I'm depressed, you can't ever say that word. It doesn't actually... exist in my language, believe me, I've tried to... translate it..., it's not there..., it's that bad"*<br>- P6, Black African/British Black African, radio presenter |
|  | Education on mental health awareness and support | *"mental health isn't really highly regarded in our community... so, we're trying to do workshops... to kind of bring awareness of it"*<br>- P7, Asian/British Asian, community service provider |
| Financial impact | Financial strain | *"during coronavirus, ... people are... struggling because... the things gone up. Prices gone high, and their earning gone down. Some people are struggling paying rent..., struggling in their jobs. For example, taxi drivers."*<br>- P5, Asian/British Asian, mosque imam |
|  | Recourse to public funds | *"What the Government did which... has impacted... people of African Caribbean origin, is that... they had a furlough scheme... which they told everyone... would end in October... Then at the very last minute... announced that they're gonna [going to]... put the furlough scheme back. But the effect was, by that time,... there was a lot of unnecessary redundancies... Black young people are twice... as likely to be unemployed than their White counterparts. So, they would have been affected by those sort of decisions"*<br>- P16, Black/African/Caribbean/British Black, community service provider |
| Physical health impact | Fear of compromised safety in seeking medical help | *"I know people who won't go outside right now because they're afraid of catching the virus because... if you're BME, you're more likely to [die] ... They don't wanna [want to] go to the doctors... Some say they're not going to hospital because, again, they were dying in hospital. They left home... not that sick, and by the time they get there, they die... So they're fearful of going to hospital... It's across... the community"*<br>- P11, Black/African/Caribbean/British Black, church leader |
|  | Implications of lockdown on current and future health | *"Within the African Caribbean community, one of the conditions... is sickle cell anaemia and thalassemia... Because of the lockdown,... they've been restricted in terms of the treatment that they can get... Men and prostate cancer,... it's very high in the African... community. If you're not able to get a check-up,... that delay can be fatal, because in African Caribbean men, ... prostate cancer tends to be... a stronger strain... So now,... there's a potential that people out there have either passed away or moved into stage four when, if services had been available, they wouldn't have... So... there will be... some deaths that are unnecessary, really, because of this lockdown"*<br>- P16, Black/African/Caribbean/British Black, community service provider |
| Social impact | Restricted cultural and religious gatherings | *"When somebody dies in the community, we get hundreds of people, it's not about 15 or 30, it's hundreds... I think a lot of our cultural norms,... it's like it's been eradicated, or just taken away... To all of a sudden restrict the amount of people who can attend church, and then when you're there, ... we can't worship like... we used to, because we're quite vocal, we're very expressive and all that's been... taken away"* |
|  |  | - P18, Black/African/Caribbean/British Black, community service provider |
|  | Fear of implications from self-isolation | *"My community are poor, most of them are... on benefits,... some of them are... working... low paid jobs, like cleaners... So...if a person is already worried if they can put food on the table,... they don't think more about the risk of what's happening out there... And... there's a lot of... single parents in Birmingham... What will I do to feed my children if the lockdown happens and... I lost my job? ... It's very difficult when you're thinking about feeding your own children and about your health, so you... may go to work... maybe not feeling well yourself... They were even hiding that they have the COVID-19 symptoms,... because they cannot afford to stay home."*<br>- P6, Black African/British Black African, radio presenter<br>*"people are not... willing to say, oh I've got symptoms, cuz [because] of fear that people are not gonna [going to] come near them, but they would talk about other people, other stories"*<br>- P7, Asian/British Asian, community service provider |
|  | Reinforced structural racism and prejudice | *"The heightened awareness of the fact that it's impacting BME [BAME] communities has given rise to...justifying hate... and we've seen some of this manifested online towards our community."*<br>- P1, Asian/British Asian, community service provider<br>*"Our people have lost a lot of family members through this COVID,... People in our culture, in our community, felt unappreciated, uncared for, lack of value placed on them, who they are, and what they've accomplished. And even how they've been treated historically, it's now coming back stronger because they're now saying... we were asked to come to this country years ago, we weren't coming here by ourselves,... doing other people's work, come and help rebuild the country. And now you've done all that, they're treating you as though you're nothing but a slave, you're just...an intruder in this country."*<br>- P11, Black/African/Caribbean/British Black, church leader |

**Table 3.** Understanding of risk and disparity in health outcomes for COVID-19.

| Thematic subtheme | Thematic Code | Illustrative quotes |
| --- | --- | --- |
| Acceptance, scepticism and ignorance |  | *"Some of the people that are from back home and they've come to this country, they have the mentality that, some, not all, some took it really seriously, but some thought if you were gonna [going to] get the virus, you were gonna [going to] get it, and they were quite reluctant to obey the rules."*<br>- P7, Asian/ British Asian, community service provider<br>*"It's a linkage [the disparity], whether it's true or not,... especially when you then compare it to India, ... whether the numbers are wrong or not, accurately counted or whatever,... but the margins cannot be that much. ...It can't be that India... underreport such less numbers compared to the UK or mainland Europe,... that it has somehow... not affected Indians ... in India to the same degree or same level as those in the UK."*<br>- P13, Asian/British Asian, temple manager |
| Perceptions around ethnic, cultural, societal and environmental contexts as risk factors | Multi-generational living | *"living with extended people, they said that was a risk of infection spreading... because the White British public,... they're quite limited to...their... literal families that they're living with, whereas we have extended families."*<br>- P7, Asian/British Asian, community service provider |
|  | Education level and employment | *"When you look at... hospitals and health establishments, the workers there are mainly from the BAME communities, so... they would be more at risk... because we're the ones... who are on the front line... to protect people... from COVID."*<br>- P18, Black/African/Caribbean/British Black, community service provider |
|  | Neighbourhood deprivation or affluency | *"The community... believe that... the Government is not doing enough in time, and sense that there's a lot of deaths that have occurred amongst our community,... because they're the poorer sectors, ... those who live in the... poor housing area"*<br>- P11, Black/African/ Caribbean/ British Black, church leader |
|  | Community collectivism | *"I think for lots of communities..., they see your family, our family as being one family, almost. So they don't differentiate and think, well actually, it's households"*<br>- P14, various BAME groups, community service provider |
|  | Poor health and lifestyle and lack of access to healthy food | *"Our community, the food is not healthy,... they don't eat balanced diet..., they don't have physical activities. And... they have different kind of... diseases like diabetic, heart...problems...and...cholesterol, and...because of this they...understand that... we are more vulnerable... compared to the White community."*<br>- P5, Asian/British Asian, mosque imam |
|  | Cultural hierarchy | *"We... stay with our grandparents, with our parents..., it's just a part of our culture.... The teenagers... understand the risk,... but what can they do? Who's in charge of the house? The grandparents or the parents are in charge...They may come to them and say, look, mummy or daddy you cannot go and visit that person who passed away..., because of COVID-19. They [in charge] say, oh, you are becoming Western, why you not understanding... it's our culture? That's our family... Young people don't have... the authority... The adults cannot understand... what's the Government saying, what's the Public Health England saying. So, they don't know, they think ... they're [young people] exaggerating, they're becoming Western, why are* |
|  |  | *they telling us not to go? So... young people understands but... our older ones... don't,... and that's where the risk lies because if they get the infection, young people gets infection because they live together."*<br>- P6, Black African/British Black African, radio presenter |

**Table 4.**
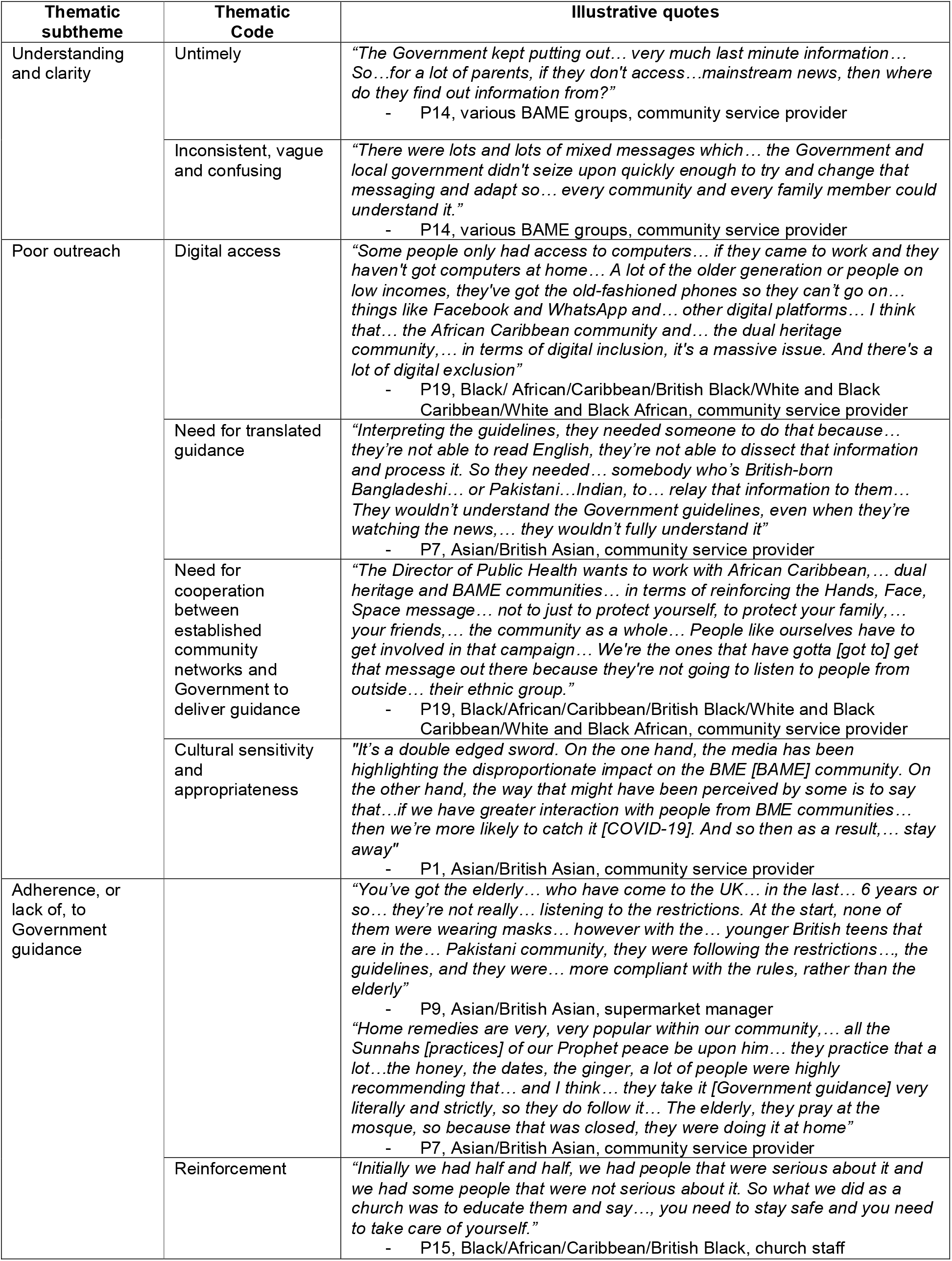
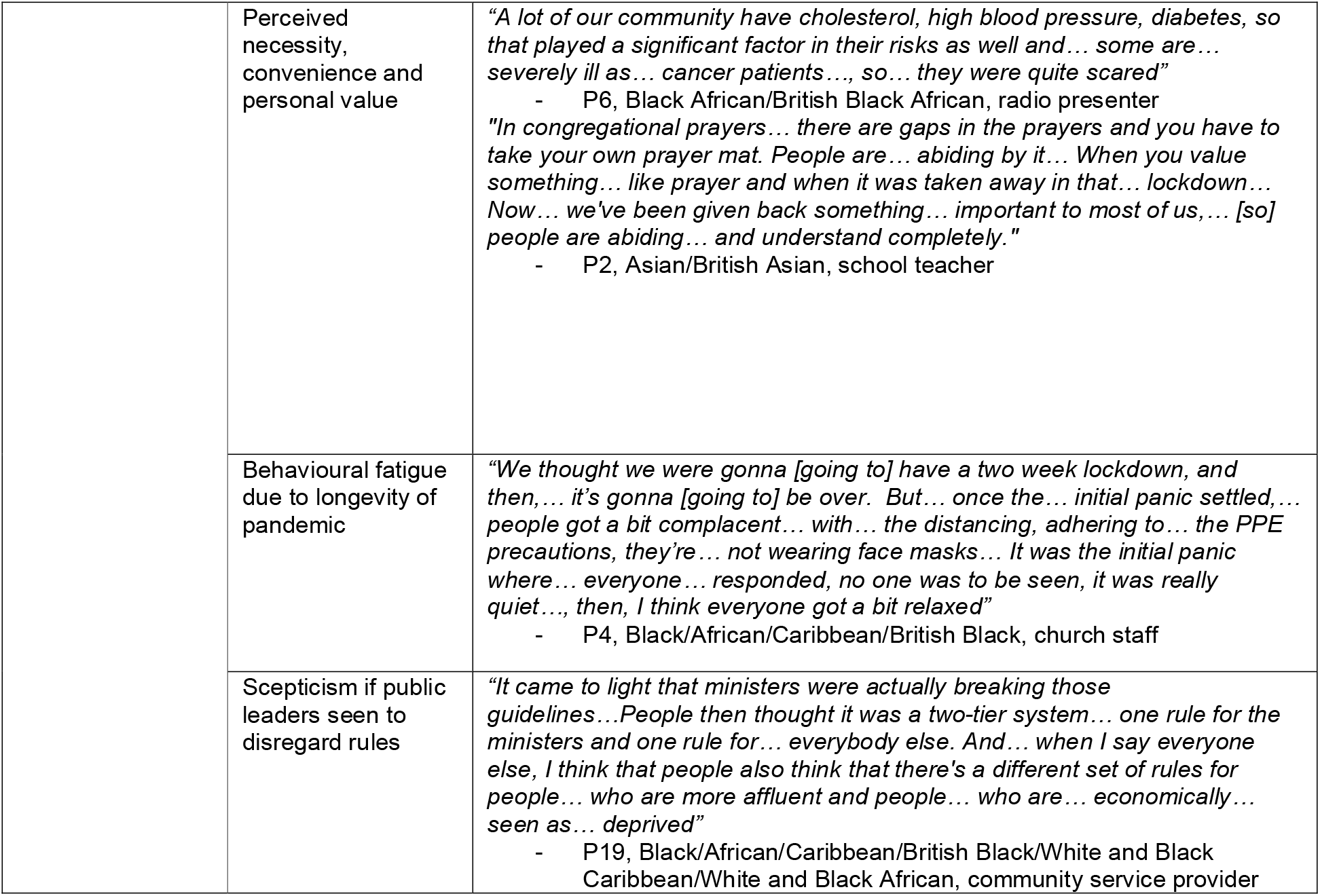
Perception, understanding and adherence to Government guidance in relation to COVID-19.

| Thematic subtheme | Thematic Code | Illustrative quotes |
| --- | --- | --- |
| Understanding and clarity | Untimely | *"The Government kept putting out... very much last minute information... So...for a lot of parents, if they don't access...mainstream news, then where do they find out information from?"*<br>- P14, various BAME groups, community service provider |
|  | Inconsistent, vague and confusing | *"There were lots and lots of mixed messages which... the Government and local government didn't seize upon quickly enough to try and change that messaging and adapt so... every community and every family member could understand it."*<br>- P14, various BAME groups, community service provider |
| Poor outreach | Digital access | *"Some people only had access to computers... if they came to work and they haven't got computers at home... A lot of the older generation or people on low incomes, they've got the old-fashioned phones so they can't go on... things like Facebook and WhatsApp and... other digital platforms... I think that... the African Caribbean community and... the dual heritage community,... in terms of digital inclusion, it's a massive issue. And there's a lot of digital exclusion"*<br>- P19, Black/ African/Caribbean/British Black/White and Black Caribbean/White and Black African, community service provider |
|  | Need for translated guidance | *"Interpreting the guidelines, they needed someone to do that because... they're not able to read English, they're not able to dissect that information and process it. So they needed... somebody who's British-born Bangladeshi... or Pakistani...Indian, to... relay that information to them... They wouldn't understand the Government guidelines, even when they're watching the news,... they wouldn't fully understand it"*<br>- P7, Asian/British Asian, community service provider |
|  | Need for cooperation between established community networks and Government to deliver guidance | *"The Director of Public Health wants to work with African Caribbean,... dual heritage and BAME communities... in terms of reinforcing the Hands, Face, Space message... not to just to protect yourself, to protect your family,... your friends,... the community as a whole... People like ourselves have to get involved in that campaign... We're the ones that have gotta [got to] get that message out there because they're not going to listen to people from outside... their ethnic group."*<br>- P19, Black/African/Caribbean/British Black/White and Black Caribbean/White and Black African, community service provider |
|  | Cultural sensitivity and appropriateness | *"It's a double edged sword. On the one hand, the media has been highlighting the disproportionate impact on the BME [BAME] community. On the other hand, the way that might have been perceived by some is to say that...if we have greater interaction with people from BME communities... then we're more likely to catch it [COVID-19]. And so then as a result,... stay away"*<br>- P1, Asian/British Asian, community service provider |
| Adherence, or lack of, to Government guidance |  | *"You've got the elderly... who have come to the UK... in the last... 6 years or so... they're not really... listening to the restrictions. At the start, none of them were wearing masks... however with the... younger British teens that are in the... Pakistani community, they were following the restrictions..., the guidelines, and they were... more compliant with the rules, rather than the elderly"*<br>- P9, Asian/British Asian, supermarket manager<br>*"Home remedies are very, very popular within our community,... all the Sunnahs [practices] of our Prophet peace be upon him... they practice that a lot...the honey, the dates, the ginger, a lot of people were highly recommending that... and I think... they take it [Government guidance] very literally and strictly, so they do follow it... The elderly, they pray at the mosque, so because that was closed, they were doing it at home"*<br>- P7, Asian/British Asian, community service provider |
|  | Reinforcement | *"Initially we had half and half, we had people that were serious about it and we had some people that were not serious about it. So what we did as a church was to educate them and say..., you need to stay safe and you need to take care of yourself."*<br>- P15, Black/African/Caribbean/British Black, church staff |

|  |  |
| --- | --- |
| Perceived necessity, convenience and personal value | *"A lot of our community have cholesterol, high blood pressure, diabetes, so that played a significant factor in their risks as well and... some are... severely ill as... cancer patients..., so... they were quite scared"*<br>- P6, Black African/British Black African, radio presenter<br>*"In congregational prayers... there are gaps in the prayers and you have to take your own prayer mat. People are... abiding by it... When you value something... like prayer and when it was taken away in that... lockdown... Now... we've been given back something... important to most of us,... [so] people are abiding... and understand completely."*<br>- P2, Asian/British Asian, school teacher |
| Behavioural fatigue due to longevity of pandemic | *"We thought we were gonna [going to] have a two week lockdown, and then,... it's gonna [going to] be over. But... once the... initial panic settled,... people got a bit complacent... with... the distancing, adhering to... the PPE precautions, they're... not wearing face masks... It was the initial panic where... everyone... responded, no one was to be seen, it was really quiet..., then, I think everyone got a bit relaxed"*<br>- P4, Black/African/Caribbean/British Black, church staff |
| Scepticism if public leaders seen to disregard rules | *"It came to light that ministers were actually breaking those guidelines... People then thought it was a two-tier system... one rule for the ministers and one rule for... everybody else. And... when I say everyone else, I think that people also think that there's a different set of rules for people... who are more affluent and people... who are... economically... seen as... deprived"*<br>- P19, Black/African/Caribbean/British Black/White and Black Caribbean/White and Black African, community service provider |

**Table 5.** Accessibility and use of community services, and other points of support, during the pandemic.

| Thematic subtheme | Thematic code | Illustrative quotes |
| --- | --- | --- |
| Accessibility and lack of resources |  | *"The wider community, they need more support. And I would [suggest]... that every faith group be given resources they need to get the people that are qualified... If the right resources are put in there, people will find more easy to... go to temples, the mosque, synagogue and the church and talk to priest, or a minister, or a qualified counsellor from that group who can deal with them based on... their faith and know who they are as a community."*<br>- P11, Black/African/Caribbean/British Black, church staff |
| Points of support | Family and friends | *"The people that speak English in the household were responsible for the shopping, whether it be online or actually attending the actual shops and getting things"*<br>- P7, Asian/British Asian, community service provider |
|  | Places of worship and religious leaders | *"The food bank over the past six months has... been inundated with people from... the community... We started off a drop-off service as well during the first lockdown... We also have a counselling service as a listening service..., people, they just, they wanna [want to] talk"*<br>- P10, various BAME groups, mosque imam |
|  | Established community networks and centres | *"We've got networks..., a lot of community organisations across the borough that's working together. So we share information and information's gone out in different languages and... we've got community champions... in the area"*<br>- P18, Black/African/Caribbean/British Black, community service provider |
|  | Healthcare professionals | *"9 of 10 people I talk to are having problems to get through to their GPs... The doctor calls [but they] don't understand what the doctor is saying... Before the pandemic, people used to take, like, okay, I'll take my friend with me... who says [English] maybe better than me... But on the phone, they can't... So if somebody doesn't speak English, or doesn't have... anyone at home... who can speak English... what will they do?"*<br>- P6, Black African/British Black African, radio presenter |

### A) Perceived impact of COVID-19 and lockdown on wellbeing

#### Mental health impact and psychological wellbeing

This was widely expressed by all participants, with feelings of anxiety being exacerbated by social isolation appearing to culminate in emotional fatigue, owing to the longevity of the pandemic. Participants alluded to the close knit nature of the BAME communities and the pandemic and the lockdown had an immense impact on their social wellbeing. Some also reported that women within families may have suffered a notable impact due to additional strain from domestic responsibilities during lockdown (table 2).

Whilst participants differed in their perspectives on whether adequate mental health support was available during and after the first national lockdown, all recognised that further education within their respective communities was required to break the taboo that presented a barrier to expressing emotion, and encourage those who require support to actively seek it. Participants of Asian and Black African communities described that mental health was still deemed a taboo within the communities and many had to suffer in isolation (table 2).

#### Financial impact

The subsequent financial impact from the national lockdown was an overriding concern for BAME communities, particularly for those on lower incomes and in self-employment. Financial insecurity as a result of the pandemic was especially difficult for families, as several participants mentioned that many in their communities were reliant on Government welfare (table 2).

#### Physical health impact

Current and future physical health was a concern expressed by many participants (table 2). They recognised that their communities are often predisposed to certain chronic conditions which may result in worse outcomes, due to the current treatment prioritisation of COVID-19. Examples described related to the impact on the ongoing treatment of patients with sickle cell anaemia, thalassemia and prostate cancer that were deemed to be highly prevalent amongst Black African communities. Therefore, those with underlying conditions were taking extra precautions to maintain optimum health, should they be affected by COVID-19.

Leaders mentioned their community’s fear of being admitted to hospital due to the worry of potentially dying away from loved ones. Mistrust in the health services appeared to be propagated by social media and prior negative experiences in healthcare; this stopped many from self-reporting if they had symptoms (table 2).

#### Social impact

Some participants reported an increase in targeted online hate crime, perhaps due to media representation of their community during this pandemic (table 2). The high prevalence of COVID-19 in BAME communities and the media representations have resurfaced historic structural biases against their community, with some commenting strongly on the perceived discrimination. Similarly, leaders noted the perceived lack of support for their respective BAME groups regarding COVID-19 and its effects.

Participants also described the taboo within their own communities meant many were suffering in isolation. Many would also be unwilling to report symptoms if it meant that they would have to self-isolate, due to consequent financial loss and, sometimes, stigmatisation from fellow community members (table 2).

Leaders expressed their communities’ struggles with restrictions on social distancing and lockdown measures that had greatly impacted on what would otherwise be high-volume cultural and religious gatherings, from festivities to funerals (table 2).

### B) Understanding of risk and disparity in health outcomes for COVID-19

#### Acceptance, scepticism and ignorance

Participants spoke of varying degrees of acceptance, with most describing their communities accepting their increased risk of transmission, infection and worse health outcomes compared to the White British population. However, scepticism of the virus’s impact was expressed by the participants, particularly due to the disproportionate death toll in the UK compared to their native country (table 3). All mentioned that the greatest health impact was on those who were elderly and vulnerable within their community, including those with underlying health conditions and disabilities (table 3).

#### Perceptions around ethnic, cultural, societal and environmental contexts as risk factors

Neighbourhood deprivation was commonly described, with many commenting on poor lifestyles and socioeconomic status fuelling the disparity (table 3). This was linked to education level and employment nature, especially for those in the service sector with high public exposure. Participants discussed multigenerational living contributing towards the high prevalence. For younger generations who were conscious of the risk, it was reported that some may have struggled to communicate this to elders within their family and wider community due to cultural hierarchies (table 3).

### C) Perception, understanding and adherence to Government guidance in relation to COVID-19

#### Understanding and clarity

Almost all participants stated that Government guidance was inconsistent and lacked clarity. The subsequent effect was worsened for community members who were not fluent in English, with translated guidance lacking in forms that could be accessed and understood by all. This, combined with the community’s close-knit nature, was reported to have perhaps influenced their interpretation of social distancing guidelines between households (table 4).

#### Poor outreach

Digital exclusivity exacerbated weak understanding, with leaders mentioning that the pandemic revealed the lack of digital access for their most deprived (table 4). Participants described how they took on the role of disseminating Government guidance; a common sentiment was that of wonder to how else their community would have received such information without their input. Some mentioned that media representation and comments by Government officials made their community feel marginalised, as it sometimes appeared to be targeted directly towards particular ethnic groups (table 4).

#### Adherence, or lack of, to Government guidance

Participants deemed that communities in more deprived areas appearing to have weaker uptake of guidelines. Other risk avoidance behaviours included the use of home remedies, which were perceived to have great benefit and were linked to cultural and religious practices (table 4).

Facilitators affecting adherence including personal value, such as being able to resume congregational worship. Reinforcement from local authority and trusted community and religious leaders encouraged adherence. In contrast, the simplicity of some guidelines (wearing face masks and avoiding non-essential travel) appeared to make some question the necessity of abiding by them, especially when Government officials were also seen to publicly break the rules without consequence (table 4).

### D) Accessibility and use of community services, and other points of support, during the pandemic

#### Accessibility and lack of resources

Some mentioned their community had restricted or no access to culturally-appropriate service providers and places of worship that they would normally frequent. Many commented on inadequate access to General Practitioners, a healthcare professional their community holds in high regard and would usually turn to (table 5).

#### Points of support

All commented on their community’s feeling of servitude towards each other during the pandemic (table 5). Family and friends were reported to be the immediate support network. As lockdown restrictions eased, community centres and places of worship were stated to have responded to their local’s needs by launching services, such as food banks and befriending projects, which were not previously required (table 5).

## DISCUSSION

The aim of this study was to investigate the impact of COVID-19 pandemic on BAME communities. The findings reveal the inequalities as experienced by leaders of diverse BAME communities. Participants alluded to the disparities in infection rates and outcomes to historical and structural differences. Many community members experienced racism and stigmatisation during the pandemic. Cultural and social practices within the communities led to many suffering on their own as discussion of mental health was still deemed a taboo within many BAME communities.

Participants identified their community members faced barriers to adherence to government guidelines. Lack of English proficiency particularly contributed to this. Despite translated documents, there was often an issue of illiteracy in the native language for some community members, with a number of participants mentioning the need for interpreters to verbally deliver guidance. It is known that deliverance of Government guidelines was markedly affected by digital exclusion. In 2019, only 10.6% of White ethnic groups in the West Midlands were found to be “internet non-users”, whilst the collective percentage for all other ethnic groups (as included in our study) was reported at 39.9%.^21^ In addition, some participants described that lack of access to sanitizers and masks during the early phase of the pandemic led to further difficulties in adherence for the most disadvantaged in their community.

Stigmatisation of mental health has frequently been documented within BAME communities; this perceived barrier to seeking support from peers or professionals were described by our study participants. Our findings also revealed a mental health impact from the lockdown particularly on women with familial responsibilities in BAME communities. This also extended to the lack of adequate bereavement support. Many communities could not grieve as usual due to lockdown restrictions. For instance, the Muslim community initially could not perform burial rites within 24 hours, an otherwise-expected practice. This was quickly ameliorated by emergency Government legislation which respected the community’s wishes.^22^ Such practice from authority level should be implemented for other minority ethnic groups regarding the particular challenges their communities face during this pandemic.

Participants described overcrowding due to multi-generational living as a risk factor for the observed disparities and contributor to weak adherence of social distancing guidelines. Many alluded to poor housing conditions. The ONS have reported that only 2% of White British households are overcrowded, compared to 10% for other ethnic groups.^23^ Participants also commented on the nature of employment their community members typically hold, including key worker roles during the pandemic (such as transport operatives and hospital porters) overexposed them to the virus.^24^ High prevalence of such employment amongst BAME communities have been linked to poorer education levels associated with historic structural biases and systemic inequality. ^24^This is reiterated by research stating that such factors interplay with ethnicity, resulting in poor health for minority ethnic groups.^25^

### Strength and limitations

To our knowledge, this was the first study to investigate the understanding of risk and impact of COVID-19 using the perspectives of BAME community leaders in England. An extensive variety of community leaders were recruited through an intensive search of BAME community organisations, businesses and places of worship. Thus, key informants could share the experiences of COVID-19 pandemic on their BAME community through the study’s qualitative design. The interviewing-researcher’s own BAME origin may have also allowed participants to openly discuss sensitive issues, thus eliciting detailed perspectives.

Duplicate analysis of interviews provided rigour, and data saturation was assumed after 19 participants since no new themes had emerged. However, these findings are not representative of all BAME groups. For example, we could not recruit anyone from East Asian communities. Leaders not of BAME origin themselves but who could speak on behalf on BAME communities (such as councillors of White ethnicity representing a West Midlands constituency with a high ethnic demographic) were approached, but we did not receive response from anyone available to participate. Moreover, our methodology’s use of Government-standardised ethnic grouping was very broad, but our results indicated that the experiences of this pandemic varied hugely across different ethnicities that would be classed together. For instance, the Somali diaspora in inner-city Birmingham had very different understanding and experiences of COVID-19 compared to the Caribbean community in outskirt boroughs.

Although not an aim of this study, there is weakness in its lack of generalisability as it was limited to a geographical region within England. However, this region represents the second highest proportion of BAME populations within England.^26^

### Implications for practice and research

Further work needs to be urgently undertaken during this ongoing pandemic to improve adherence to the Government guidelines within BAME communities and mitigate the disproportionate impact of COVID-19. This includes increasing outreach and providing logistical and financial support at a grassroots level to the most vulnerable in already-marginalised communities. Public health guidance must be produced in different languages and dialects through accessible media, not only on how to stay safe from COVID-19 but to also challenge myths propagated by social media. Public health campaigns should incorporate nuances that BAME communities can resonate with, such as the perceived benefit of home remedies, to deliver targeted but culturally sensitive interventions. All aforementioned recommendations should be implemented with cooperation between health services and trusted community networks, religious leaders and local stakeholders. This approach would be beneficial in other global or national public health interventions and any future pandemics, should they occur.

Future research could investigate which intra-demographic characteristics within a certain BAME community affect their perceptions and impact of COVID-19, and to what extent. This includes factors identified by this study, such as cultural hierarchies. The collective addressing of all non-White demographics into one overarching BAME category should also be questioned and adapted, as this study has demonstrated that one solution will not encompass the needs of all BAME communities. Perspectives of other population groups who are likely to face multiple social disadvantage during the time of pandemic such as the homeless populations,^27–29^ refugees^30^ and single people living^31^ needs to be researched.

## CONCLUSION

BAME community leader participants of this study alluded to historical and structural differences for the observed disparities in COVID-19 morbidity and mortality. In addition, cultural and social practices within the communities led to many suffering on their own as discussion of mental health was still deemed a taboo within many communities. Racial discriminations added to their worries during the pandemic. Reinforcement from trusted community and religious leaders encouraged adherence to government guidelines. Points of support such as food banks were vital in ensuring essential supplies during the pandemic. Government agencies and public health bodies need to integrate with the community, and community leaders to penetrate the key messages and deliver targeted yet sensitive public health advice which incorporates cultural and religious practices. Addressing route cause of disparities is imperative to mitigate current and future pandemics. These must be addressed by using appropriate and targeted public health interventions. Such interventions should be emboldened by the governmental sphere foremost, then perpetuated by local authority in collaboration with community leaders. Ultimately, all strategies must be guided by BAME communities themselves in order to successfully meet their needs.

## Supporting information

Electronic supplement materials 1 and 2

## Data Availability

All relevant data pertaining to this manuscript are presented in the manuscript and the electronic supplementary materials.

## Acknowledgments

We are grateful to every participant for their time and support in this study, without whom this research could not have been conducted.

## Author contributions

FM, VP and DA co-designed the study. FM conducted all interviews and transcribed the data. FM and VP conducted the analysis in duplicate to which DA and KK added their input through expert comments. FM led the write up of the manuscript to which all authors contributed through editing and expert comments. All authors agree to the final version of the manuscript.

## Checklist

This work follows COREQ recommendations and is reported as per the COREQ checklist. (Electronic Supplement 3)

