## Supplementary material for "Impact of COVID-19 pandemic on Black, Asian and Minority Ethnic (BAME) communities: a qualitative study on the perspectives of BAME community leaders": Electronic supplement materials 1 and 2

**Electronic supplementary material 1**

Ethnic groups of interest for study participation as a Black, Asian or Minority Ethnic community

| **Broad ethnic group** | **Ethnicities included** |
| --- | --- |
| Black/African/Caribbean/Black British | African; Caribbean and any other Black/ African/ Caribbean background |
| Asian/Asian British | Indian; Pakistani; Bangladeshi; Chinese and any other Asian background |
| Other | Arab and any other ethnic group |
| Mixed/Multiple | White and Black Caribbean; White and Black African; White and Asian and any other mixed/multiple ethnic background |

Adapated from: GOV.UK. List of ethnic groups (no date). Available at: https://www.ethnicity-facts-figures.service.gov.uk/style-guide/ethnic-groups [Accessed January 8, 2021].

**Electronic supplementary material 2: Interview Topic Guide**

**Understanding of acquiring COVID-19 risk and disparity in health outcomes**

1) How has the experience of your community been so far with the COVID-19 pandemic?
2) How does your community perceive the risks of transmission and infection?
   a. Do all members of your community understand its risk equally, or do some members understand the risk more than others?

[Probes: different age groups, gender, education level, employment status, income level, those living with families, those with health conditions]

3) How does your community perceive the risks when compared to white British population?

[Probes: higher rates of infection, hospitalisation, ICU admission, and death]

**Community beliefs and perspectives**

4) In your opinion, what do your community believe are the reasons for their higher risk of COVID-19 incidence and their disparity in health outcomes?

[Probes: genetics, vitamin D deficiency, housing quality, cultural gatherings, congregational worship, employment nature, health practices (smoking and alcohol intake, sedentary lifestyle), socio-economic status, community collectivism]

5) Are there any risk avoidance behaviours that your community have adopted?
   a. Please describe and explain these behaviours

[Probes: social distancing, not meeting in households, wearing masks in public spaces, avoiding public transport]

6) Do your community feel comfortable discussing the risks of COVID-19?

[Probes: any perceived stigmas, taboo]

7) How do you perceive the impact of COVID-19 on the a) physical, b) social and c) mental wellbeing of your community?

[Probes: exercise, healthy eating, personal recreation, financial security for: housing, food, utilities]

a. Are some members of your community affected more than others? Please explain

[Probes: different age groups, gender, education level, employment status, income level, those living with families, those with health conditions]

b. What is your understanding of how COVID-19 has impacted those within your community with disabilities?

[Probes: deaf, blind, learning difficulty, those requiring additional care]

c. Where do your community members turn to for support? Why?

[Probes: healthcare workers, religious leaders]

d. Do your community believe they had access to adequate mental health support during and after the national lockdown? Why?
e. Could the support have been improved, and how?

8) What are the main concerns of your community relating to COVID-19 and the subsequent impact on their wellbeing?

[Probes: impact on: education, employment, management of current health conditions and accessing health services]

9) Is there anything else you would like to discuss about your community’s perspective of COVID-19, and the impact it has had on them?

**Understanding and implementation of government-issued guidance**

10) The Government has issued many communication materials to raise public awareness on how to stay safe and reduce the infection rate of COVID-19. How does your community find in interpreting the information and guidelines?

[Probes: IT literacy, access to digital media, those who do not socialise outside of their households, those who cannot speak English fluently, use of over-the-counter medicines]

11) What is your understanding of how COVID-19 has impacted those who cannot speak or understand English fluently?

[Probes: understanding of public health information]

12) How is government advice about social distancing, washing and hygiene practices being perceived and implemented within your community? Please explain.

[Probes: staying 2m apart, not mixing with other households, limiting travel, restricting shopping movements]

a. What would make your community members more or less likely to adopt government guidance on staying safe and reducing the infection rate?

13) How do you perceive the impact of social distancing and lockdown restrictions on your community?

[Probes: on family, economy, education, health]

14) Who do your community members trust and approach for information on COVID-19?
15) Does your community understand where to reach out for help and services when they have symptoms?

[Probes: if self-isolating]

16) Are there any factors stopping your community members from seeking advice about COVID-19, ranging from diagnosis to treatment and staying safe?

[Probes: immigration fears, fear of racism, mistrust in health services, fear of implications from being told to self-isolate]

17) In your opinion, what could be done to lift these barriers?
    a. Can healthcare workers provide additional support to your community? If yes, how so?

**Final comments**

18) Are there any final comments you wish to make about your community’s perspectives of COVID-19 and how it has impacted them?
